# Correlation of long-term care facility vaccination practices between seasons and resident types

**DOI:** 10.1101/2021.10.05.21264257

**Authors:** Emily T O’Neill, Elliott Bosco, Erin Persico, Joe B Silva, Melissa R Riester, Patience Moyo, Robertus van Aalst, Matt M Loiacono, Ayman Chit, Stefan Gravenstein, Andrew R Zullo

## Abstract

**Objectives:** Influenza vaccination varies widely across LTCFs due to staff behaviors, LTCF practices, and patient factors. It is unclear how seasonal LTCF vaccination varies between cohabitating but distinct short-stay and long-stay residents. Thus, we assessed the correlation of LTCF vaccination between these populations and across seasons.

**Design:** National retrospective cohort using Medicare and Minimum Data Set (MDS) data.

**Setting and Participants:** U.S. LTCFs. Short-stay and long-stay Medicare-enrolled residents age ≥65 in U.S. LTCFs from a source population of residents during October 1^st^-March 31^st^ in 2013-2014 (3,042,881 residents; 15,683 LTCFs) and 2014-2015 (3,143,174, residents; 15,667 LTCFs).

**Methods:** MDS-assessed influenza vaccination was the outcome. Pearson correlation coefficients were estimated to assess seasonal correlations between short-stay and long-stay resident vaccination within LTCFs.

**Results:** The median proportion of short-stay residents vaccinated across LTCFs was 70.4% (IQR, 50.0-82.7%) in 2013-2014 and 69.6% (IQR, 50.0-81.6%) in 2014-2015. The median proportion of long-stay residents vaccinated across LTCFs was 85.5% (IQR, 78.0-90.9%) in 2013-2014 and 84.6% (IQR, 76.6-90.3%) in 2014-2015. Within LTCFs, there was a moderate correlation between short-stay and long-stay vaccination in 2013-2014 (*r=*0.50, 95%CI: 0.49-0.51) and 2014-2015 (*r=*0.53, 95%CI: 0.51-0.54). Across seasons, there was a moderate correlation for LTCFs with short-stay residents (*r=*0.54, 95%CI: 0.53-0.55) and a strong correlation for those with long-stay residents (*r=*0.68, 95%CI: 0.67-0.69).

**Conclusion and Implications:** In LTCFs with inconsistent influenza vaccination across seasons or between populations, targeted vaccination protocols for all residents, regardless of stay type, may improve successful vaccination in this vulnerable patient population.

**Brief summary:** In LTCFs, influenza vaccination was moderately correlated across all residents, but varied across seasons. Inconsistent vaccination among cohabitating residents may increase infection risk in LTCFs.

## INTRODUCTION

Ninety percent of influenza-related deaths occur in patients ≥65 years old.^1^ Older adults residing in long-term care facilities (LTCFs) are particularly vulnerable to seasonal influenza infections. Ease of inter-resident transmissibility within LTCFs in combination with increased frailty and multimorbidity, contributes to the high burden of influenza in this population.^2, 3^ Prevention of outbreaks are primarily dependent upon diligent staff hygiene and proper infection control practices, in addition to staff and resident vaccination adherence.^4^ While several U.S. Food and Drug Administration approved agents for chemoprophylaxis are recommended in the event of an influenza outbreak, vaccination is a cornerstone of prevention.^5^

The Centers for Medicare & Medicaid Services (CMS) require participating LTCFs to assess all residents and provide influenza vaccine if indicated. At the beginning of 2020, the CMS Office of Disease Prevention and Health Promotion established a target threshold of 90% for seasonal influenza vaccination among adults in LTCFs.^6^ Despite this goal, variation in influenza vaccination across U.S. LTCFs exists, with one study reporting U.S. LTCF vaccination ranging from 50-89.7% in 2014-2015.^7^ Differences in vaccination coverage across LTCFs is likely multifactorial and the result of structural inequalities, LTCF quality, and adequacy of staffing levels or training.^8-12^ An understudied yet growing area of importance for infection prevention is the interplay between the commonly cohabitating short-stay and long-stay residents.^8, 13, 14^ Though a CMS quality measure for “Influenza Vaccination Assessment and Provision” is reported on Nursing Home Compare for each population within LTCFs, seasonal differences in vaccination of short-stay and long-stay residents are not systematically explored.^15^

Determining LTCF vaccination consistency, which can be defined as similar vaccine administration across different seasons and resident types, is necessary to identify barriers to achieving vaccination goals among these vulnerable populations. Stratifying by both season and resident type is an important first-step to understanding the etiologies of LTCF vaccination coverage. For instance, major inconsistencies between resident types in a given LTCF might suggest lack of parity in vaccination policies for each population. Inconsistencies across seasons for one population might reflect vaccine supply or storage barriers within the LTCF.

Thus, in this study we 1) estimated the proportion of short-stay and long-stay residents receiving influenza vaccines in U.S. LTCFs, 2) assessed the correlation of LTCF vaccination between these populations in a given season, and 3) assessed the correlation of LTCF vaccination for each population across influenza seasons. We hypothesize that a strong correlation (Pearson correlation coefficient *r* > 0.60) would exist for each population within and across seasons due to similar vaccination policies and access to resources within LTCFs.

## METHODS

### Data Sources and Study Design

Minimum Data Set (MDS) version 3.0 LTCF resident clinical assessments were linked to the Medicare Master Beneficiary Summary File (MBSF), Certification And Survey Provider Enhanced Reports System (CASPER), and LTCFocus facility data using unique identifiers for all LTCF residents enrolled in Medicare. These data have been previously described.^8^ This was a retrospective cohort study derived from a source population of U.S. LTCF residents with stays during October 1^st^-March 31^st^ in 2013-2014 (N=3,042,881 residents; 15,683 LTCFs) and 2014-2015 (N= 3,143,174 residents; 15,667 LTCFs).

### Study Population

Eligible LTCF residents were classified as short-stay with a total stay of <100 days in the same LTCF, or long-stay with a total stay of ≥100 consecutive days and no more than 10 days outside of the LTCF. The date of LTCF entry and 100^th^ day in the facility were considered index dates for short- and long-stay residents, respectively. Those included in the study population had six months of continuous enrollment in Medicare Part A before index and were ≥65 years old at index. We excluded: i) residents of hospital-based LTCFs because these differ markedly from most LTCFs in their structure, and ii) residents of LTCFs with missing CASPER data. The institutional review board at Brown University approved the study protocol. Due to the use of de-identified administrative data, informed consent was not required.

### LTCF Influenza Vaccination Measures

MDS assessments from October 1^st^ through June 30^th^ in each season were used to determine resident vaccination status, as is common in CMS quality measures.^15^ Receipt of influenza vaccination was assessed through a previously published algorithm that utilizes vaccine responses from the MDS.^16^ Residents were considered vaccinated if any MDS assessment during the study period indicated the resident received influenza vaccination at that facility or outside of the facility, prior to entry. The proportion vaccinated for each resident group, short-stay and long-stay, was calculated by dividing the number of residents vaccinated within a facility by the total number of residents meeting inclusion criteria in that facility. Additionally, we calculated the “Appropriately Assessed and Provided Vaccination” (AAPV) measure which is reported on Nursing Home Compare and is a composite of vaccinated, refused, and contraindicated MDS responses.^15^ Overall, we calculated four vaccination outcomes: vaccinated, refused, contraindicated, or AAPV.

### Covariates

Demographic variables such as age, sex, and race/ethnicity were obtained from the MBSF. LTCF-level variables, including staffing and care quality measures, were obtained from CASPER and LTCFocus data.

### Statistical Analyses

Median LTCF-level vaccination measures were calculated for each population and each season along with the interquartile range (IQR). Pearson correlation coefficients were estimated to assess the correlation between an individual LTCF’s proportion of residents vaccinated: i) comparing short-stay and long-stay residents within a given season, and ii) either the proportion of short-stay or long-stay residents vaccinated across seasons. Correlation coefficients were estimated using LTCFs with short-stay and long-stay residents across the 2013-2014 and 2014-2015 influenza season. Data were analyzed using SAS version 9.4 (SAS Institute, Inc., Cary, NC) and R version 3.6 (R Foundation for Statistical Computing, Vienna, Austria).

## RESULTS

We included 14,116 LTCFs with short-stay residents and 14,473 LTCFs with long-stay residents in the 2013-2014 season, and 14,203 LTCFs with short-stay and 14,444 with long-stay residents in the 2014-2015 season (**Supplementary Table S1, Supplementary Figure S1**). Characteristics of LTCFs with short-stay versus long-stay residents were similar across seasons.

The median proportion of short-stay residents vaccinated in LTCFs was 70.4% (IQR: 50.0-82.7%) in 2013-2014 and 69.6% (IQR: 50.0-81.6%) in 2014-2015 (**Table 1**). The short-stay AAPV measure was a median of 91.3% (IQR: 79.2-98.8%) in 2013-2014 and 90.4% (IQR: 78.6-97.2%) in 2014-2015. The median proportion of long-stay LTCF residents vaccinated was 85.5% (IQR: 78.0-90.9%) in 2013-2014 and 84.6% (IQR: 76.6-90.3%) in 2014-2015 (**Table 1**). The long-stay AAPV measure was a median of 97.6.% (IQR: 94.5-100%) in 2013-2014 and 97.2% (IQR:93.8-99.1%) in 2014-2015. Across both seasons and for both populations, vaccination refusals were a larger component response for the AAPV measure than contraindication to vaccination. Median proportions of LTCF residents refusing influenza vaccination were higher among short-stay than long-stay residents.

**Table 1:**
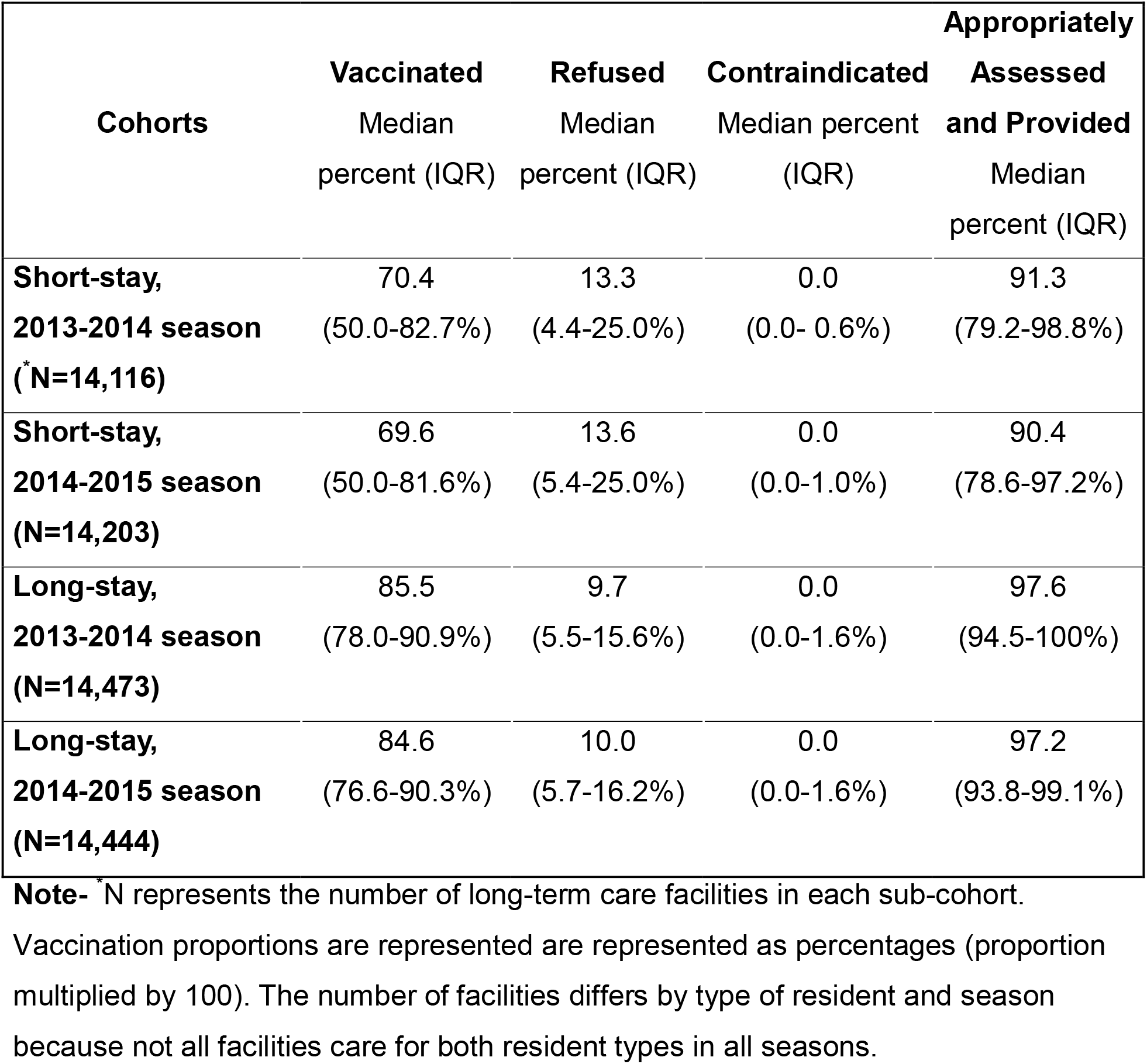
Long-term care facility vaccination measures by type of resident and influenza season.

Within LTCFs, there was a moderate correlation between the proportion of short-stay and long-stay vaccinated in 2013-2014 (*r*=0.50, 95% CI: 0.49-0.51) and 2014-2015 (*r*=0.53, 0.51-0.54) (**Table 2 and Figure 1A-D**). There was a moderate correlation across seasons for short-stay residents (*r*=0.54, 0.53-0.55) and a strong correlation for long-stay residents (*r*=0.68 0.67-0.69) (**Table 2 and Figure 1A-D**).

**Table 2:**
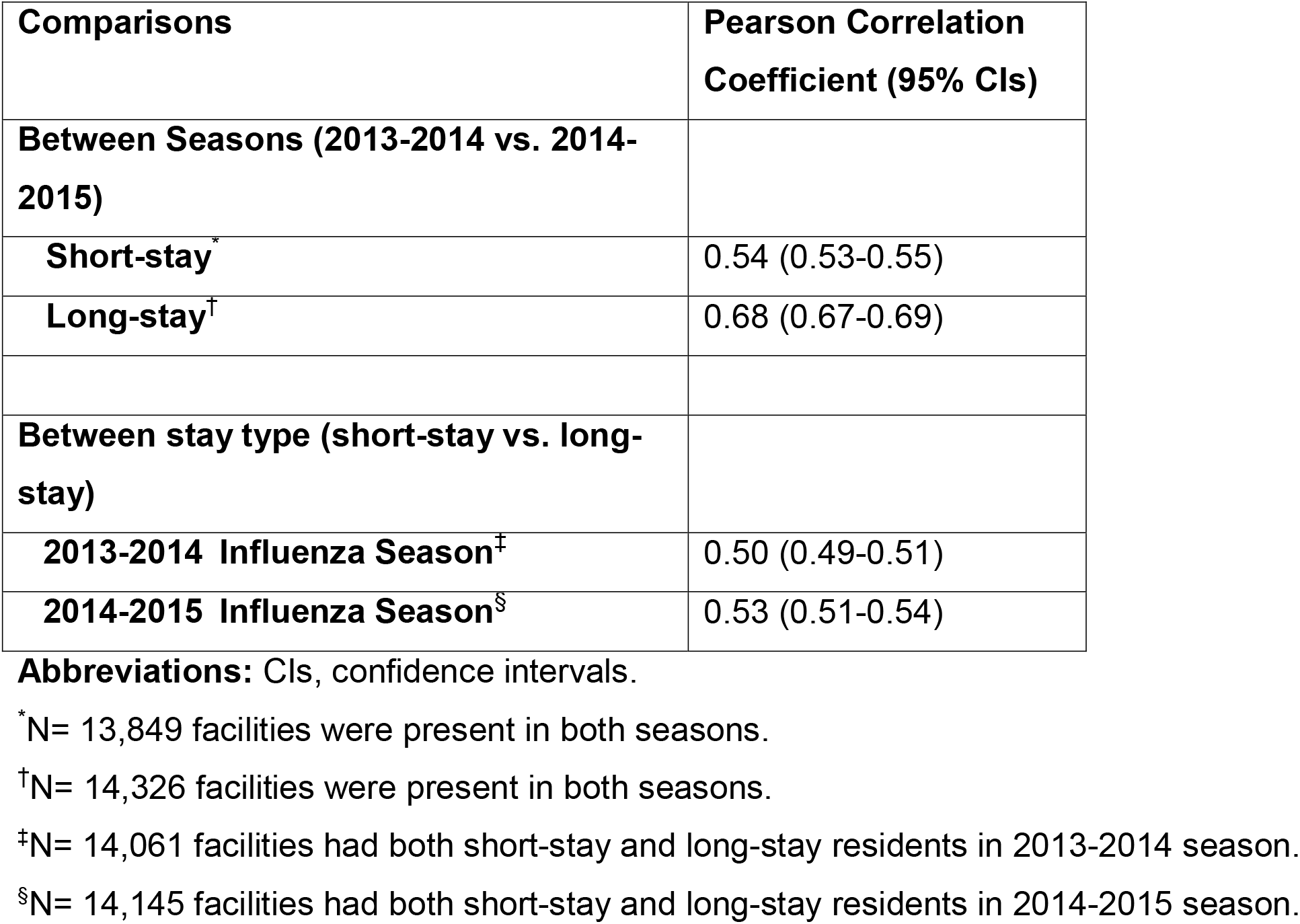
Correlations of LTCF proportion of residents receiving influenza vaccine by season and stay type.

| **Comparisons** | **Pearson Correlation Coefficient (95% CIs)** |
| --- | --- |
| **Between Seasons (2013-2014 vs. 2014-2015)** |  |
| **Short-stay<sup>\*</sup>** | 0.54 (0.53-0.55) |
| **Long-stay<sup>†</sup>** | 0.68 (0.67-0.69) |
| **Between stay type (short-stay vs. long-stay)** |  |
| **2013-2014 Influenza Season<sup>‡</sup>** | 0.50 (0.49-0.51) |
| **2014-2015 Influenza Season<sup>§</sup>** | 0.53 (0.51-0.54) |

**Abbreviations:** CIs, confidence intervals.
<sup>\*</sup>N= 13,849 facilities were present in both seasons.
<sup>†</sup>N= 14,326 facilities were present in both seasons.
<sup>‡</sup>N= 14,061 facilities had both short-stay and long-stay residents in 2013-2014 season.
<sup>§</sup>N= 14,145 facilities had both short-stay and long-stay residents in 2014-2015 season.

**Figure 1A-D:** Plots of the within-LTCF-level vaccination proportion correlations between short-stay and long-stay residents and across influenza seasons. A; The proportion of short-stay residents vaccinated versus the proportion of long-stay residents vaccinated in the 2013-2014 season for LTCFs with both populations. B; The proportion of short-stay residents vaccinated versus the proportion of long-stay residents vaccinated in the 2014-2015 season for LTCFs with both populations. C; The proportion of short-stay residents vaccinated in the 2013-2014 season versus the proportion of short-stay residents vaccinated in the 2014-2015 for LTCFs present in both seasons. D; The proportion of long-stay residents vaccinated in the 2013-2014 season versus the proportion of long-stay residents vaccinated in 2014-2015 for LTCFs present in both seasons. Each black dot represents a single LTCF. The blue line represents the line of best fit.

## DISCUSSION

Our study assessed LTCF influenza vaccination among short-stay and long-stay residents in the 2013-2014 and 2014-2015 influenza seasons. Over the course of the study period, the proportion of short-stay residents vaccinated was lower than the proportion of long-stay residents vaccinated. Long-stay resident influenza vaccination was consistent across seasons, indicated by a strong correlation, though vaccination of short-stay residents was not consistent. Within each season, vaccination of both populations residing in the same LTCF was not consistent, indicated by a moderate correlation. Compared to LTCF long-stay vaccination measures, proportions of residents refusing vaccination were higher in short-stay populations and AAPV measures were lower. These findings suggest that short-stay residents may be both assessed and offered vaccination less frequently, while also having higher refusal rates.

Patient factors that differ across short-stay and long-stay residents may drive the observed differences in vaccination rates. Short-stay residents typically spend a short time in LTCFs while recovering from acute illness, while long-stay residents are typically older with physical and cognitive impairment that could prevent independent living.^8,17^ With shorter duration of illness among short-stay patients, it is possible that lower vaccination rates may be attributed to the LTCFs inability to obtain seasonal influenza vaccine in a timely manner prior to discharge. Further, acute illness among short-stay residents may capture the attention of providers, taking preference over ordering preventive measures such as influenza vaccination. Among long-stay residents, increased frailty and decreased autonomy over healthcare decisions may contribute to the higher vaccination coverage. Cognitive impairment may further decrease decision-making capacity of long-stay residents, enabling practitioners to obtain vaccination consent more easily from healthcare decision-makers.^18^

Non-patient factors such as vaccination procurement and standing orders could lead to the discrepancies observed between short-stay and long-stay populations. Additionally, these factors may impact vaccination rates by affecting LTCFs ability to obtain the vaccine and effectively administer it. Influenza vaccine stock fluctuates across seasons, leaving some providers and facilities with limited resources, and ultimately impacting which patients receive priority for vaccination. Fluctuations in purchasing costs across health care settings further disrupt vaccine procurement. For example, LTCFs that purchase limited vaccine quantities may pay higher prices due to missed bulk ordering rebates.^19-21^ Differences in vaccination programs among LTCFs such as the use of standing orders, vaccination consent and refusal protocols, provider reminders, and frequent review and audit of administration policies may catalyze differences across LTCFs.^18,20^ Minimizing these identified barriers may increase rates of influenza vaccination among this vulnerable population.

Our study builds on existing literature assessing policy “spill-over” within LTCFs where protocols aimed at one resident type impact non-targeted residents.^22^ We hypothesized that correlation between resident types and across seasons would be strong due to positive “spill-over” effects of vaccination policies within the LTCF. That is, the custodial care provided to long-stay residents and processes to administer seasonal influenza vaccine would extend to short-stay residents within an LTCF. From this, we assumed that LTCFs with high long-stay resident vaccination proportions would have similarly high vaccination of short-stay residents. Unfortunately, our study reveals varying levels of consistency across resident types, with a larger proportion of long-stay residents consistently vaccinated compared to their short-stay counterparts. Susceptibility of older adults to influenza and other respiratory pathogens such as SARS-CoV-2 underlies the need for consistently high levels of vaccination coverage among LTCF residents.^1, 23^ Further research assessing vaccination variation among resident types and across seasons is needed to develop tailored vaccination protocols that captures all residents.

The current study is not without limitations. We were unable to assess if the influenza vaccine was truly received by the patient prior to LTCF entry due to the use of MDS data. The MDS influenza vaccination questions are self-reported measures and may be inaccurate, however they have been found to be valid measures of influenza vaccination.^24^ Future studies should integrate vaccination data across settings of care to assess this. We also did not assess the health status of LTCF residents; these factors may vary across LTCFs and determine the resident’s likelihood of vaccination. Further, differences in vaccination protocols or ability to obtain vaccine may influence LTCF vaccination but were not assessed due to lack of available data. Lastly, we chose to focus on two influenza seasons during non-pandemic (e.g., SARS-CoV-2 or 2009 influenza H1N1) time periods to provide a non-pandemic measure of LTCF vaccine administration. These findings may not extrapolate well to vaccination behaviors during pandemics or over a longer time-period.

## CONCLUSIONS AND IMPLICATIONS

Our research identified inconsistencies in vaccination rates between long-stay and short-stay residents across the 2013-2014 and 2014-2015 influenza seasons. LTCFs that vaccinate inconsistently across residents may benefit from standard protocols independent of resident type, while variation across season may indicate the need for increased vaccine stocks or staffing adherence. We hope these findings can further inform LTCFs and public health agencies to develop tailored vaccination policies, and ultimately improve influenza vaccination receipt in this vulnerable patient population.

## Data Availability

Minimum Data Set (MDS) version 3.0 LTCF resident clinical assessments were linked to the Medicare Master Beneficiary Summary File (MBSF), Certification And Survey Provider Enhanced Reports System (CASPER), and LTCFocus facility data using unique identifiers for all LTCF residents enrolled in Medicare. These data have been previously described

## Acknowledgements

E.O. and E.B. participated in conceiving the study, data collection, data analysis, interpretation of the data, writing and critical revision of the manuscript for important intellectual content, and final approval of the manuscript submitted. S.G. and A.R.Z. participated in conceiving the study, data collection, interpretation of the data, critical revision of the manuscript for important intellectual content, and final approval of the manuscript submitted. E.P., J.S, M.R.R., P.M, R.V.A, M.L., and A.C. participated in interpreting results, providing critical revisions, and final approval of the manuscript submitted. The authors would like to acknowledge Melissa Eliot and Jessica Ogarek for their help with data management and analysis. Employees of the sponsor (R.V.A., M.L., and A.C.) participated in interpretation of the data, review of the manuscript, and final approval of the submitted manuscript. The study sponsor was not responsible for analyzing the data or preparing the initial manuscript draft.

