## Supplemental Documents for "Correlation of long-term care facility vaccination practices between seasons and resident types"

**Supplementary Material**

**Supplementary Figure S1.** Sample selection diagram.

**Supplementary Table S1.** LTCF demographics represented as means (standard deviation) or counts (percentage- represented by %).

**Supplementary Figure S1.** Sample selection diagram.

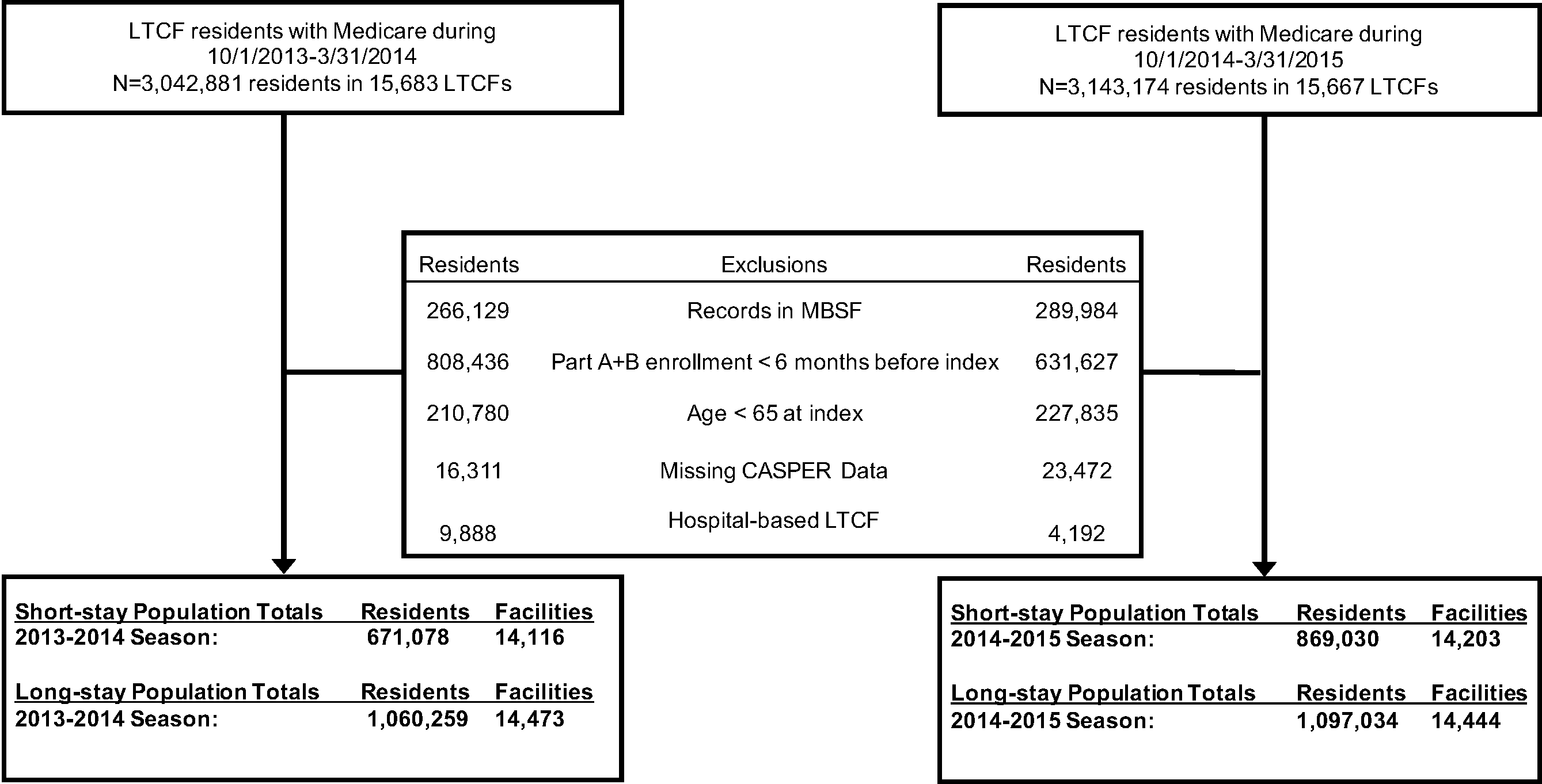

**Abbreviations**: LTCFs, long-term care facilities; MBSF, Medicare master beneficiary summary file.

**Supplementary Table S1.** LTCF demographics represented as means (standard deviation) or counts (percentage- represented by %).

| **LTCF Characteristics** | **Short-stay,**  **2013-2014**  **(**^*^**N=14,116)** | **Long-stay,**  **2013-2014**  **(N=14,473)** | **Short-stay,**  **2014-2015**  **(N=14,203)** | **Long-stay,**  **2014-2015**  **(N=14,444)** |
| --- | --- | --- | --- | --- |
| **Demographics** |  |  |  |  |
| **Age, years, mean (SD)** | 79.7 (3.6) | 83.8 (3.5) | 80.2 (3.5) | 83.7 (3.5) |
| **Race/ethnicity, %, mean (SD)** |  |  |  |  |
| **Non-Hispanic White** | 81.0 (22.6) | 80.8 (22.8) | 80.0 (23.0) | 79.8 (23.2) |
| **Non-Hispanic Black** | 10.5 (16.9) | 10.5 (17.0) | 10.6 (17.0) | 10.7 (17.0) |
| **Hispanic** | 4.1 (10.3) | 4.2 (10.5) | 4.1 (10.4) | 4.2 (10.4) |
| **Structural Characteristics** |  |  |  |  |
| **Average daily census, mean (SD)** | 89.6 (52.3) | 88.9 (52.2) | 88.7 (51.7) | 89.6 (52.3) |
| **Occupancy rate, %, mean (SD)** | 82.6 (14.0) | 82.6 (14.0) | 82.4 (14.1) | 82.6 (14.1) |
| **Number of beds, mean (SD)** | 110.7 (59.2) | 109.8 (59.1) | 110.2 (59.0) | 109.8 (59.0) |
| **Urban, n (%)** | 9,940 (70.4) | 10,162 (70.2) | 10,016 (70.5) | 10,162 (70.2) |
| **For-profit status, n (%)** | 10,369 (73.5) | 10,593 (73.2) | 10,465 (73.7) | 10,601 (73.4) |
| **Chain ownership, n (%)** | 8,085 (57.3) | 8,192 (56.6) | 8,211 (57.8) | 8,271 (57.3) |
| **Resident primary insurance, %, mean (SD)** |  |  |  |  |
| **Medicare** | 15.3 (13.2) | 14.7 (12.7) | 15.1 (13.4) | 14.6 (12.9) |
| **Medicaid** | 60.3 (21.7) | 61.0 (21.7) | 60.1 (21.9) | 60.8 (21.8) |
| **Acuity Index,^a^ mean (SD)** | 12.0 (1.5) | 12.0 (1.6) | 12.1 (1.3) | 12.1 (1.4) |
| **Admissions/bed, mean (SD)** | 2.0 (1.8) | 1.9 (1.7) | 2.0 (1.8) | 2.1 (1.6) |
| **Staffing** |  |  |  |  |
| **Total Nursing HPRD, mean (SD)** | 3.9 (1.4) | 3.9 (1.3) | 4.0 (1.3) | 4.0 (1.3) |
| **CNA HPRD, mean (SD)** | 2.4 (0.8) | 2.4 (0.8) | 2.4 (0.8) | 2.4 (0.7) |
| **RN HPRD, mean (SD)** | 0.5 (0.4) | 0.4 (0.3) | 0.5 (0.4) | 0.5 (0.3) |
| **LPN HPRD, mean (SD)** | 0.8 (0.4) | 0.8 (0.4) | 0.8 (0.4) | 0.8 (0.4) |
| **Ratio of RN/RN+LPN, mean (SD)** | 0.3 (0.2) | 0.3 (0.2) | 0.4 (0.2) | 0.4 (0.2) |
| **Ratio of CNA/RN+LPN, mean (SD)** | 2.0 (0.6) | 2.0 (0.6) | 1.9 (0.6) | 1.9 (0.6) |
| **Social worker on-staff hours/100 beds, mean (SD)** | 1.2 (1.1) | 1.1 (1.1) | 1.2 (1.1) | 1.1 (1.0) |
| **Quality Characteristics** |  |  |  |  |
| **Residents with bedsores, %, mean (SD)** | 6.1 (4.6) | 6.1 (4.6) | 6.2 (4.6) | 6.1 (4.6) |
| **Residents with antipsychotics, %, mean (SD)** | 22.5 (13.7) | 23.1 (14.7) | 21.6 (13.7) | 22.5 (14.6) |
| **Hospitalizations per resident-year, mean (SD)** | 1.1 (1.1) | 1.1 (1.0) | 1.1 (1.3) | 1.1 (1.3) |

**Note- ^*^**N represents the number of long-term care facilities in each sub-cohort.

**Abbreviations:** LTCF, long-term care facilities; HPRD, hours per resident day; RN, registered nurse; LPN, licensed practical nurse; CNA, certified nursing assistant; ^a^ Acuity Index is a measure of the care needed by long-term care facility residents derived from the Minimum Data Set, with a higher value indicating a greater need for care.^25^ It is calculated based on the number of residents needing various levels of activities of daily living assistance and the number of residents receiving special treatment (e.g., respiratory care, intravenous therapy, etc.).
